# Survival and 30-days hospital outcome in hospitalized COVID-19 patients in Upper Egypt: Multi-center study

**DOI:** 10.1101/2020.08.26.20180992

**Authors:** Aliae AR Mohamed Hussein, Islam Galal, Mohammed Mustafa Abdel Rasik Mohamed, Mohamed Eltaher AA Ibrahim, Shazly B Ahmed

## Abstract

**Background:** Defining the clinical features and associated outcomes of patients diagnosed with corona virus disease (COVID-19) is fundamental to improving our understanding and adequate management of this illness. **The aim of this study** was to identify the demographic data, underlying comorbidities and the mortality related factors of hospitalized patients with COVID-19 in Upper Egypt.

**Patients and methods:** 1064 cases consecutively admitted to isolation hospitals in Upper Egypt. All cases had confirmed COVID-19 infection. The electronic records of the patients were retrospectively revised and the demographic data, clinical manifestations, qSOFA score on admission and 30 days-outcome (ICU admission, death, recovery, referral or still in hospital) were analyzed. Overall cumulative survival in all patients and those > or < 50 years were calculated.

**Results:** 49.2% of the study population were males and 50.8% were females with mean age 49.4±17.8 years-old. On admission, 83.9% were stable with qSOFA score <1, 3% required non-invasive mechanical ventilation, and 2.1% required O2 therapy. Within 30 days, 203 cases (19.1%) required admission to ICU. Death was recorded in 11.7% of cases, 28.7% recovered, 40.5% referred and 19.2% were still under treatment. Determinants of ICU admission and survival in the current study were age > 50, respiratory rate > 24/minute, SaO2 < 89%, qSOFA >1 and need for O2 therapy or NIV. The cumulative survival was 75.3% with the mean survival was 28.1, and 95.2% overall survival was recorded in those aged ≤50 years.

**Conclusions:** Age older than 50 years old, those with pre-existing DM, initial qSOFA score, requirement for O_2_ therapy and NIV from the first day of hospital admission may be associated with unfavorable 30 days- in hospital outcome of COVID-19.

## Introduction

Through history, there have been plenty of pandemics however; the social response to corona virus disease (COVID-19) is unparalleled. It is assessed that almost four billion individuals are living in social segregation during this mother of all pandemics (1).

Initially described in China in December 2019, severe acute respiratory syndrome caused by corona virus 2 (SARS-CoV-2) has spread all over the world and by 18^th^ July 2020-there was an emergent figure of 13,824,739 confirmed cases and 591.666 losses reported to the WHO (2). To date, Egypt reported slightly over 82,000 confirmed COVID-19 cases with 3858 deaths (3). The new pandemic is injuring not only health organizations of several countries but also the financial prudence universal.

Defining the clinical features and associated outcomes of patients diagnosed with coronavirus disease (COVID-19) is fundamental to improving our understanding and adequate management of this illness. Several articles have recently been published, describing the clinical features and outcomes of retrospective individuals with COVID-19 (4,5,6).The findings from three previous studies suggested that older age and baseline comorbidities were associated with higher severity of disease or even death of patients with COVID-19 (7,8,9). Recently in Kuwait study, **Almazeedi et al**. found an association between several risk factors and admission to the ICU: namely, age > 50 years old, smoking, higher quick SOFA score, elevated levels of C reactive protein (CRP) and procalcitonin (PCT) levels. Furthermore, they identified a strong correlation of mortality with history of bronchial asthma, current smoking and elevated levels of PCT (10). In a previous study from China, their main hospitalized subjects were principally men with average age 56 years: 26% required ICU admission, and there was a 28% death rate (6). However, there are substantial variances between countries and Egypt in population demographical data and prevalence of comorbidities.

**The aim of this study** was to identify the demographic data, underlying comorbidities and the mortality related factors of hospitalized patients with COVID-19 in Upper Egypt.

## Patients and methods

This study included 1064 patients consecutively admitted to isolation hospitals in Upper Egypt (Aswan 67%, Luxor 19.6%, Qena 8.5% and 4.9 from Hurghada, Assiut, Sohag and Minia isolation Hospitals) during the period from 22 March to 23 June 2020; the flowchart of our cohort was demonstrated in **Figure 1**. All cases had confirmed COVID-19 infection (based on PCR, chest HRCT, CBC, CRP, Ferritin levels) and received standardized treatment protocols of Ministry of Health and Population based on disease severity (3). The electronic records of the patients were retrospectively revised and the demographic data, clinical manifestations, quick Sequential Organ Failure Assessment (qSOFA) score on admission (systolic blood pressure <100 mmHg, respiratory rate >22 breaths/min, and altered mental status, an optimum cut-off value of 1 was used to predict in-hospital mortality and qSOFA >1 were considered unstable case) (11) and 30 days-outcome (ICU admission, death, recovery, referral or still in hospital), overall cumulative survival were analyzed.

**Figure 1.**
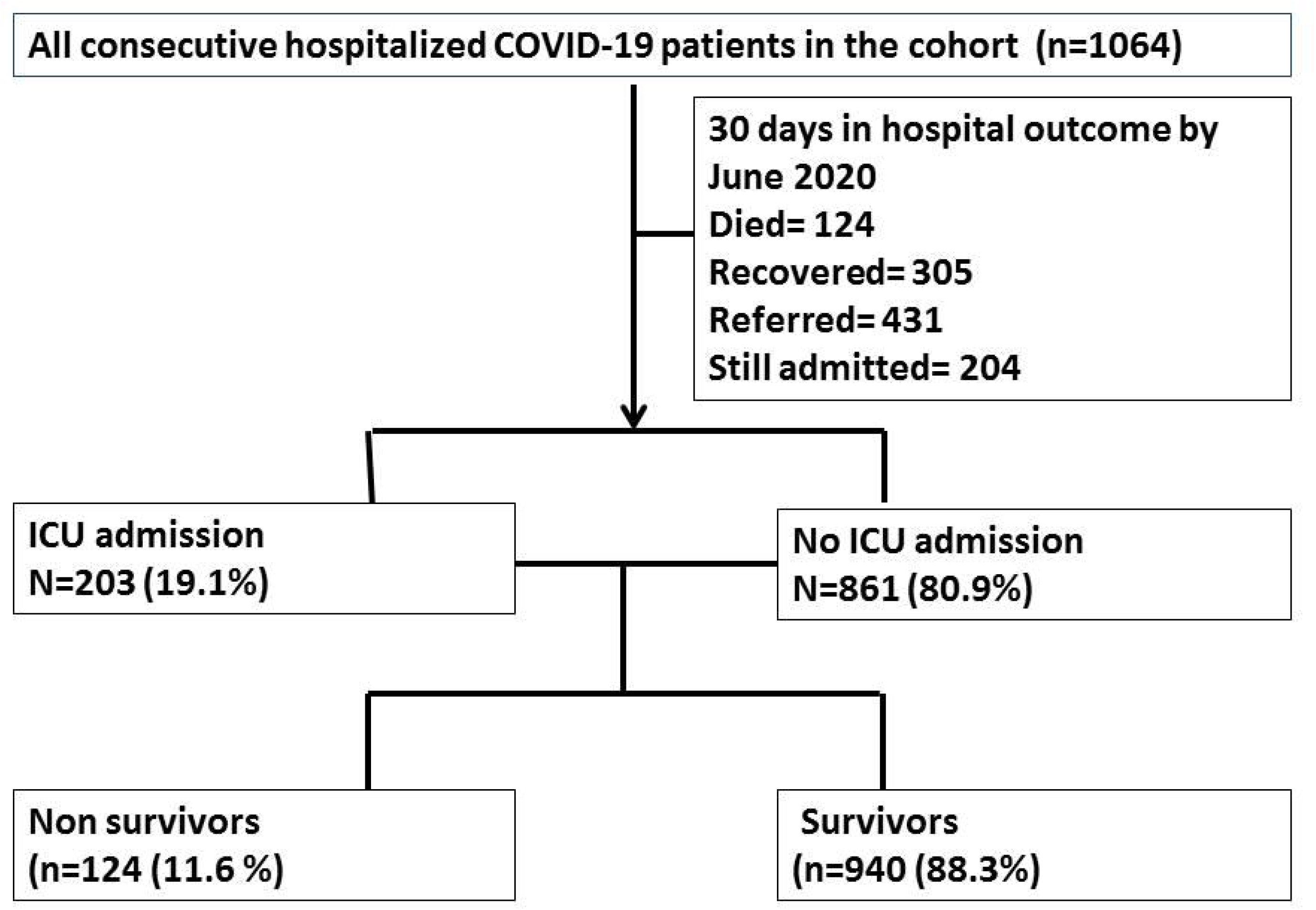
Flowchart of 30-days hospital outcome of included patients with COVID-19

### Exclusion criteria

only hospitalized confirmed cases of COVID-19 were included.

### Ethical consideration

the study was approved by ethical committee of Assiut faculty of medicine and Ministry of Population and Health Egypt.

## Statistical analysis

Data was coded and statistically analyzed using the Statistical Package of Social Science software program, version 25 (IBM SPSS 25 Statistics for windows, Armonk, NY: IBM Corp). Data was presented using range, mean, standard deviation, median and interquartile range for quantitative variables and frequency and percentage for qualitative ones. Comparison between survival & non-survival groups for qualitative variables was performed using Chi-square or Fisher’s exact tests, while for quantitative variables, the comparison was conducted using Mann Whitney test. Kaplan Meier survival analysis was conducted to calculate the cumulative overall survival rates using Log Rank test for comparison between age groups. P values less than or equal to 0.05 was considered substantially significant.

## Results

### Clinical data on admission

The current study described the characteristics of COVID-19 cohort of 1064 patients admitted in Upper Egypt Hospitals. They were 49.2% males and a 50.8% females, mean age was 49.4±17.8, median was 50 years old (range 1-95 years old). Most cases (79.3%) were contacts to positive cases, 2.8% were health care providers. On admission, 83.9% were stable with qSOFA score <1, 3% required non-invasive mechanical ventilation, and 2.1% required O2 therapy. Within 30 days, 203 cases (19.1%) required admission to ICU. Death was recorded in 11.7% of hospitalized cases, 28.7% recovered, 40.5% referred and 19.2% were still under treatment. Of 205 patients - with complete records- the mean systolic-diastolic blood pressure, diabetic status, random blood sugar, temperature, respiratory rate, pulse rate and arterial oxygen saturation were recorded in *Table 1*.

**Table 1.** Baseline descriptive data of the included cases with confirmed covid-19 admitted to hospitals (n=1064)

| Variable | Description(n=1064) |
| --- | --- |
| **Gender (n=1064)** |  |
| Male | 524 (49.2) |
| Female | 540 (50.8) |
| **Age, years (n=1061)** |  |
| Mean ± SD | 49.4 ± 17.8 |
| Median (IQR) | 50 (36 - 63) |
| **Systolic blood pressure, mmHg (n=205)** |  |
| Range | 83 - 180 |
| Mean ± SD | 122.5 ± 14.1 |
| Median (IQR) | 120 (110 - 130) |
| **Diastolic blood pressure, mmHg (n=205)** |  |
| Range | 30 - 105 |
| Mean ± SD | 75.7 ± 10.5 |
| Median (IQR) | 80 (70 - 80) |
| **Respiratory rate, breath/minute (n=205)** |  |
| Range | 15 - 47 |
| Mean ± SD | 22.2 ± 4.4 |
| Median (IQR) | 22 (20 - 24) |
| **Temperature, °C (n=205)** |  |
| Range | 36 - 38 |
| Mean ± SD | 37 ± 0.4 |
| Median (IQR) | 37 (36.9 - 37) |
| **Pulse, beat/min (n=205)** |  |
| Range | 60 - 130 |
| Mean ± SD | 89.3 ± 12.7 |
| Median (IQR) | 89 (81 - 96) |
| **SaO<sub>2</sub> (n=205)** |  |
| Range | 40 – 99 |
| Mean ± SD | 94.3 ± 7 |
| Median (IQR) | 96 (93 - 98) |
| **Diabetic Mellitus, n (%) (n=205)** | 28 (13.7) |
| **Blood sugar level of diabetic cases, mg/dl (n=28)** |  |
| Range | 98 – 449 |
| Mean ± SD | 232 ± 108.6 |
| Median (IQR) | 196.5 (144 - 294.5) |
| **Baseline condition on admission (qSOFA )(n=1064)** |  |
| Stable (qSOFA <1) | 893 (83.9) |
| Unstable (qSOFA >1) | 171 (16.1) |
| **CPAP requirement on admission (n=1064)** | 32 (3) |
| **Need for O2 therapy on admission (n=1064)** | 22 (2.1) |
| **Contact with a COVID-19 positive case** | 844 (79.3) |
| **Health care Provider** | 30 (2.8) |
| **Travelers** | 10 (0.9) |
| **Need for ICU admission ( n=1064)** | 203 (19.1) |
| **Hospital outcome in 30-days (n=1064)** |  |
| Death | 124 (11.7) |
| Recovery | 305 (28.7) |
| Referral | 431 (40.5) |
| Still admitted | 204 (19.2) |

**SD**= standard deviation, **IQR**= interquartile range (range between 25<sup>th</sup> -75<sup>th</sup> percentiles)
SaO2= oxygen saturation; CPAP=continuous positive airway pressure; ICU=intensive care unit

### Determinants of ICU admission and survival within 30 days

*In table 2*, we observed that gender was not associated with different outcome. However, 78.2% of patients aged > 50 years (median recorded age IQR) required ICU admission compared to 21.8% aged ≤ 50 years old (P=0.001). The mean age of ICU admitted patients was 60.4±14.2 vs. 46.8±17.6 (median, 62 (52-71) vs. 46 (33-61), P <0.001. The initial respiratory rate of 24.8±6.2 (median 24 (20-26) was considerably related with ICU admission as well as initial SaO_2_ of 89.7±11.4 (P< 0.001). 63.1% of patients with unstable overall condition qSOFA>1 on admission needed ICU within 30 days, moreover those who needed CPAP or O2 therapy required later ICU admission (P<0.001).

**Table 2.**
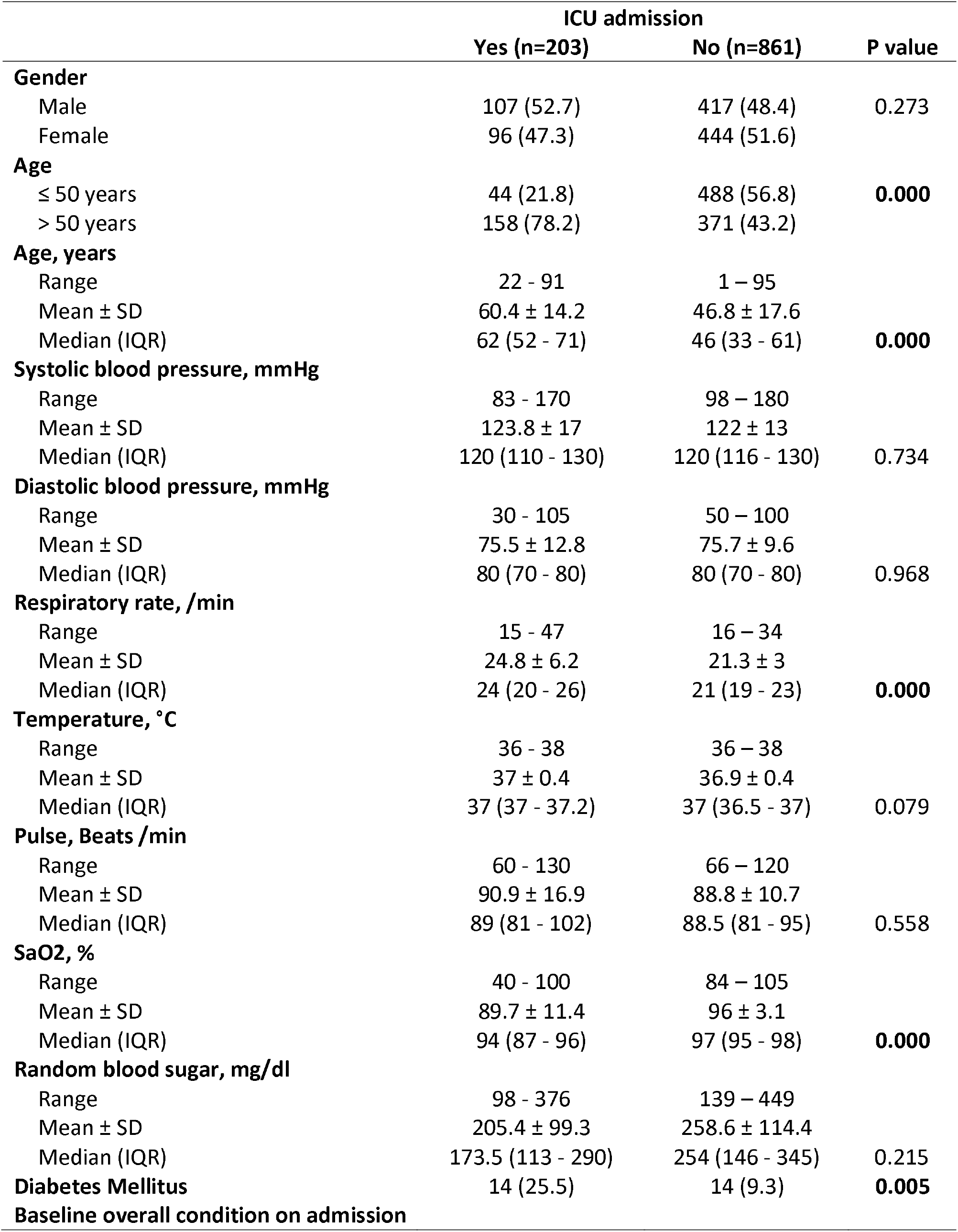
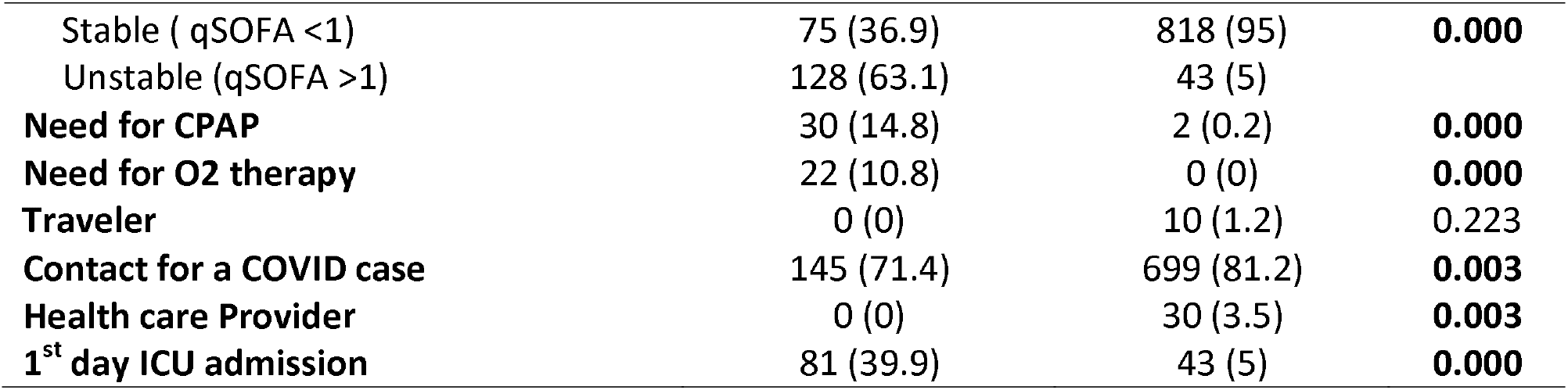
Determinants of ICU admission in hospitalized patients with COVID-19 (n=1064)

*Table 3* showed that out of 1064 patients diagnosed with COVID-19, the survival rate was 88.3% (940/1064). The survivors were considerably ≤ 50 years aged, with mean age 47.3±17.3 compared to 64.8±13.6 in non-survivors (P<0.001). They had stable baseline status on admission (95% of cases) while 100% of non survivors were unstable on admission (P<0.001). The history of admission to ICU in 1^st^ day was considerably higher in non survivors (65.3% vs. 13%, P< 0.001).

**Table 3.** Determinants of survival in studied covid-19 patients admitted to hospitals (n=1064)

|  | Outcome |  | P value |
| --- | --- | --- | --- |
|  | Non-survivors<br>(n=124) | Survivors<br>(n=940) |  |
| **Gender** |  |  |  |
| Male | 63 (50.8) | 461 (49) | 0.712 |
| Female | 61 (49.2) | 479 (51) |  |
| **Age** |  |  |  |
| ≤ 50 years | 16 (12.9) | 516 (55.1) | **0.000** |
| > 50 years | 108 (87.1) | 421 (44.9) |  |
| **Age** |  |  |  |
| Range | 20 - 91 | 1 - 95 |  |
| Mean ± SD | 64.8 ± 13.6 | 47.3 ± 17.3 |  |
| Median (IQR) | 65.5 (57.5 - 73) | 47 (34 - 61) | **0.000** |
| **Baseline condition on admission** |  |  |  |
| Stable, qSOFA <1 | 0 (0) | 893 (95) | **0.000** |
| Unstable, qSOFA <1 | 124 (100) | 47 (5) |  |
| **Need for CPAP on 1<sup>st</sup> day admission** | 5 (4) | 27 (2.9) | 0.409 |
| **Need for O2 therapy on 1<sup>st</sup> day admission** | 2 (1.6) | 20 (2.1) | 1.000 |
| **Traveler history** | 0 (0) | 10 (1.1) | 0.616 |
| **Contact with a COVID case** | 72 (58.1) | 772 (82.1) | **0.000** |
| **Health care Provider** | 0 (0) | 30 (3.2) | **0.040** |
| **1<sup>st</sup> day ICU admission** | 81 (65.3) | 122 (13) | **0.000** |

*Figure 2* showed that the overall cumulative survival in the study group is 75.3% with mean survival time (95% CI) was 28.1 (26.8-29.4). *Figure 3* demonstrated that the cumulative overall survival in patients ≤ 50 years aged was 95.2% compared to 63.0% in patients >50 years aged with mean survival time (95% CI) was 30.7 (30.1-31.4) vs. 24.9 (23.1-26.7), P<0.001.

**Figure 2.**
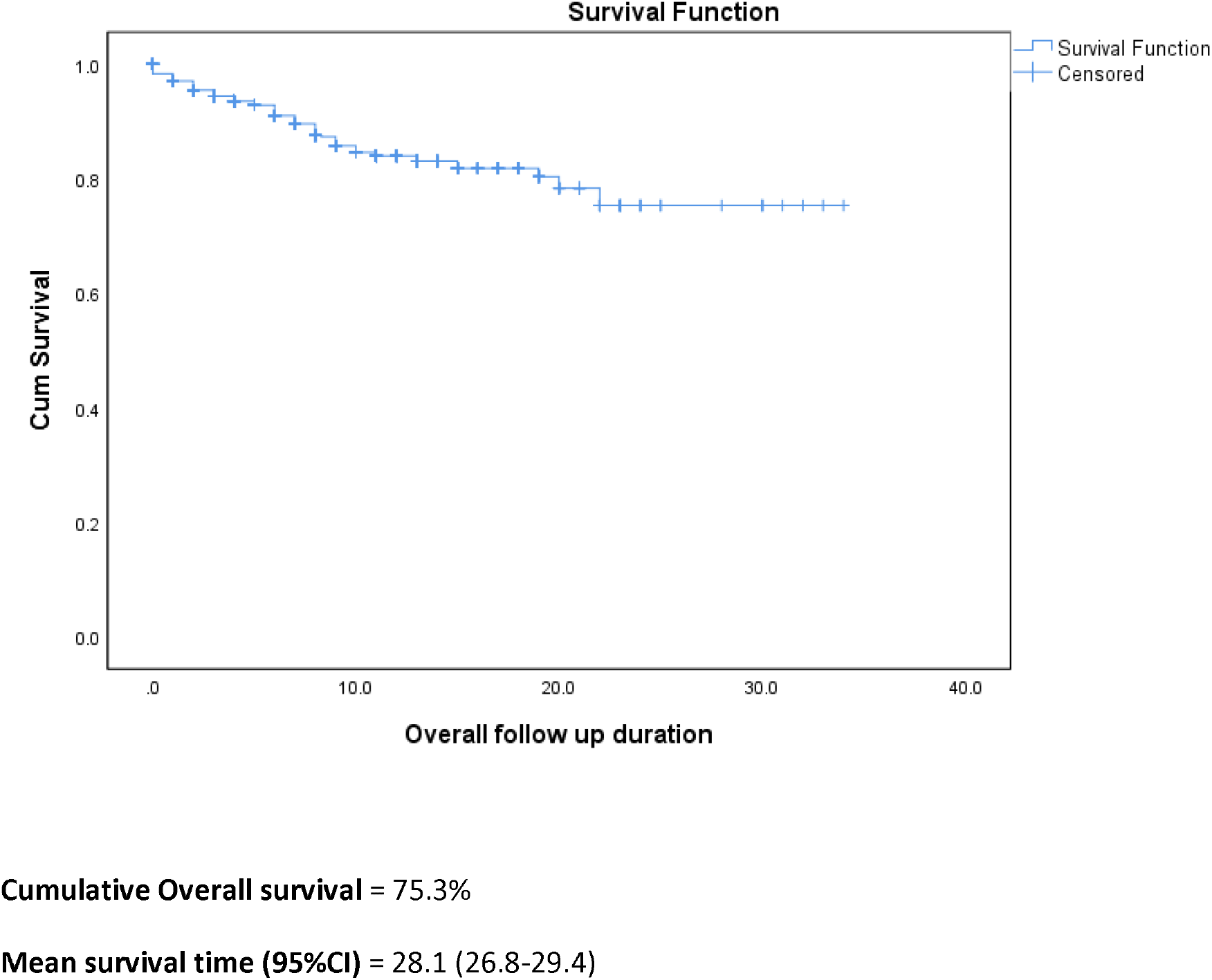
Kaplan Meier survival analysis for cumulative overall survival in Covid-19 patients admitted to hospitals

**Figure 3.**
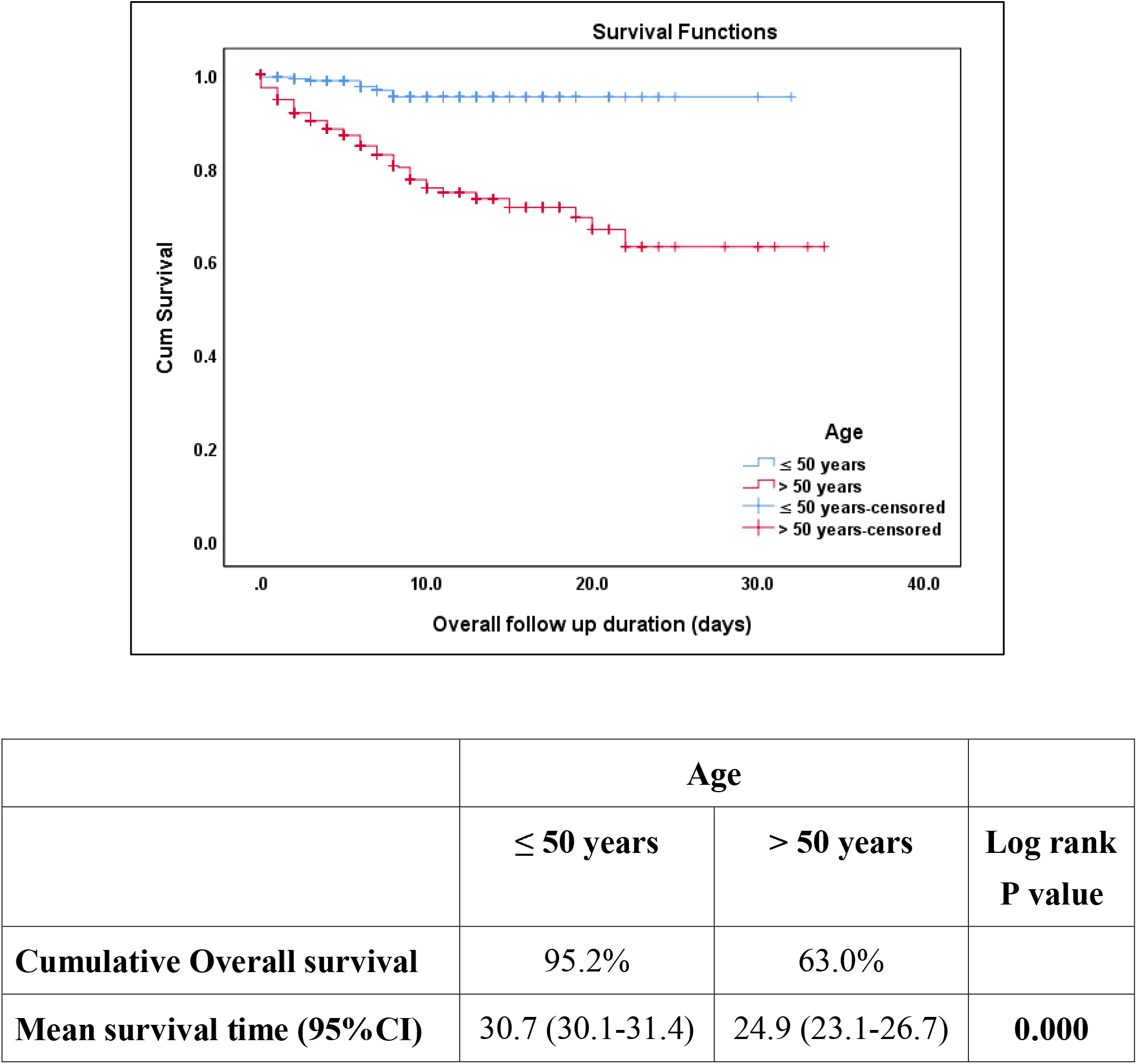
Kaplan Meier survival analysis for cumulative overall survival in patients with COVID-19 patients aged > or < 50 years old

## Discussion

The finding of this study showed that initial assessment on admission may forecast the 30 days outcome in patients hospitalized with COVID-19. Determinants of ICU admission in the current study is age > 50 years old, respiratory rate > 24/minute, SaO2 < 89%, qSOFA >1 and need for O_2_ therapy or NIV. Survival is associated with age < 50 years, initial qSOFA <1, no ICU admission in 1^st^ day. The cumulative survival is 75.3% with mean survival was 28.1, and 95.2% overall survival is recorded in those aged < 50 years aged.

Unlike many studies, the present study did not record different hospital outcome in male versus female. Similar global statistics, showing no major sex differences in the absolute number of COVID-19 confirmed cases in several countries where sex-disaggregated statistics were available (12). On the contrary, first records from China have pointed to a sex imbalance considering detected cases and the fatal outcome rate of COVID-19 (4). Richardson et al., also reported a low rate of admission for women compared to men (41.9% and 39.7%, correspondingly (5). It was suggested that usually men are involved in manual labor works and spend more time exposed to infection outdoors and this may be a contributing factor to the higher proportion of men getting the infection.

Across one meta analysis, the majority of cases were male, which supports previous studies (13,14), and a higher proportion of admitted men died than did admitted women (15). Reports addressing the sex discrepancy in COVID-19 incidence and disease course are still lacking and a thorough analysis of underlying causes is currently deficient.

Regarding the age of hospitalized patients, age seems very important contributing factor for both 30 days outcome and survival in the current study. 78.2% of patients aged > 50 years (median recorded age IQR) required ICU admission compared to 21.8% aged ≤ 50 years old (p=0.001). The mean age of ICU admitted patients was 60.4±14.2 vs. 46.8±17.6 (median, 62 (52-71) vs. 46 (33-61), P< 0.001. The survivors were extensively ≤ 50 years old, with mean age 47.3±17.3 compared to 64.8±13.6 in non survivors (P< 0.001), the cumulative overall survival in patients ≤ 50 years old was 95.2% compared to 63.0% in cases >50 years old with the mean survival time (95% CI) was 30.7 (30.1-31.4) vs. 24.9 (23.1-26.7), P< 0.001.

Since the outbreak of COVID-19, studies showed that patients in the deceased group were much older than the survivors (16), moreover univariate and multivariate logistic regression analysis concluded that age 0 65 years was a powerful forecaster of death from COVID-19 pneumonia (4, 7,9,17). In fact, in a cohort of 179 COVID-19 pneumonia patients, nobody died who was younger than 50 years, whereas 81% of the deceased patients were older than 65 years (18). A cohort of 1716 Chinese medical staff whose age was always < 65 years, only six (0.3%) died (19). There was also a strong relationship between unfortunate outcomes as ICU admission and age beyond 50 years old, a qSOFA above 0, smoking, greater CRP levels and elevated PCT levels (5, 20, 21). These data propose that the majority of patients with COVID-19 pneumonia will convalesce from the disease, mainly younger population.

In the present study, 25.5% of cases admitted to ICU were diabetics compared to 9.3% of patients not admitted to ICU, P < 0.005. The prevalence of diabetes in cases with a critical form of COVID-19 ranges from 15 to 25%, a figure which was 2 to 4-fold higher than that in non-critical cases (4). A prevalence exceeding 50% was even reported in the United States in patients admitted to ICU (7). It is reported that 72% patients of COVID-19 with comorbidities including diabetes required admission in ICU, compared to 37% of cases without comorbidities (9) and diabetic patients had a hazard ratio (HR) of 2.34 (95% CI, 1.35 to 4.05; p=0.002) for acute respiratory syndrome (ARDS) (20). Chen et al., found a considerable correlation between COVID-19 severity and diabetes (OR, 2.67, 95% CI; 1.91 to 3.74; p0.01) (8).

Interestingly, the prevalence of non-survivors was also elevated in diabetic subjects with COVID-19 and it varied from 22 to 31% in diverse studies. In a univariate analysis of 191 patients with COVID-19, Zhou et al. found that diabetes to have (an odds ratio) OR of 2.85 (95% CI, 1.35 to 6.05; p<0.001) for in-hospital mortality (6). And in a bivariate cox regression analysis, Wu et al., demonstrated a HR of 1.58 (95% CI, 0.80 to 3.13, p=0.19) for death in patients with diabetes with COVID-19 (20). In another study by Guo et al., diabetic patients died much more often than non-diabetic patients (10.8% versus 3.6%) (21).

The finding in this study did not record increase blood pressure as predictor of ICU admission and mortality. This disagree with results of a recent systematic review and meta-analysis of 14 published articles involving 4659 patients which showed that hypertension was a common underlying condition amongst all cases (39.9%), and the prevalence was substantially elevated (56.8%) in the non-survival group (15,16, 17). Hypertension confers a greater than 2.5-fold increase in the odds of death from COVID-19 in the supporting previous studies (22, 23). Further assessment is needed to explain whether hypertension alone or with other risk factors as old age, DM and cardiovascular comorbidities is a reason of worse prognosis.

Our current data demonstrated that patients in the non -survivor group were susceptible to multiple organ failure, as evaluated by qSOFA score on first day of admission. In critically ill covid-19 patients, early medical intervention to reduce mortality depends on early effective assessment. SOFA score is a good diagnostic marker and reflects the state and degree of multi-organ dysfunction. The qSOFA at admission is very useful for the prediction of mortality risk in critically ill covid-19 patients (24). Despite vaguely inferior, qSOFA is acceptable especially in the pandemic due to shortage of medical resources.. It has better-quality because it is simple, rapid and practical (25).

In one study sample, a relationship between elevated qSOFA and admission to the intensive care unit was found (10). High qSOFA and SOFA score was found to be associated with decease outcome by Zhou et al.(6). Our findings did not find a relationship between qSOFA score and death; however we did find a correlation with ICU admission. This may be due to the small number of deaths in our sample.

A mortality rate in this cohort was 11.6%. This is much lesser than other large retrospective cohort studies. Wu et al. recorded 21.9% mortality and 26.4% ICU admission (20), Zhou et al. reported 28.3% mortality and 26% ICU admission (6). Other studies recorded lower mortality rate, Guan et al., 1.4% and 5%, Richardson et al., 1.7% and 3.6% (4,5). This may imitate that these studies were one of the initial COVID-19 retrospective cohort studies so the incorporated patients may have been asymptomatic, had milder symptoms, younger age compared to studies that were published later, when health resources became more restricted and hospitalization was limited to moderate and severe cases.

Most of the limitations in this study are due to its retrospective character, for instance possible loss of data owing to omissions. In addition, we were not able to obtain the patients’ previous medical data and co-morbidities from the electronic medical records, they were recorded based on patient self-reporting and/or diagnosed using laboratory results during their admission. Moreover, during the course of pandemic, the guidelines for diagnosis, management and discharge criteria rapidly evolved ultimately, which may have implications on our results. At the end of the study, some patients remained hospitalized (19.2%) and their clinical course is still unclear.

This study is, to our knowledge, the first comprehensive study to provide data on the initial 1064 consecutive hospitalized COVID-19 cases in Upper Egypt, all admitted to isolation hospitals, undergoing the same investigations and treatment protocols. We believe it gives a different perception on the nature and clinical course of this novel disease, compared to other large retrospective cohort studies, as it included patients with a wide variety of disease severity. Future studies, directed at further characterizing risk factors for disease severity and outcomes, principally predictive scoring systems, are desired.

## Conclusions

Age older than 50 years old, those with pre-existing DM, initial high qSOFA score, requirement for O_2_ therapy and NIV from the first day of hospital admission may predict unfortunate 30 days-in hospital outcome (need for ICU and mechanical ventilation, discharge, death) among the overall population of COVID-19 cases.

## Data Availability

Available on request

## Acknowledgments

None

